# Variants in *NR6A1* as a cause for congenital renal, vertebral and uterine anomalies

**DOI:** 10.1101/2025.01.08.24319478

**Authors:** Adeline Jacquinet, Lydie Flasse, Manon Dohet, Romane Vanhaeren, Hélène Pendeville, Carol Saunders, Anna Lehman, Catherine Pienkowski, Karine Morcel, Daniel Guerrier, Vincent Bours, Bernard Peers

## Abstract

The underlying cause for renal and uterine agenesis remains unknown in many cases, whereas recurrence in some families strongly suggests the involvement of genetic factors. Here, we identify 5 affected individuals from 3 families with phenotypes including bilateral or unilateral renal agenesis/hypoplasia, along with variable congenital uterine anomalies and costovertebral defects associated with heterozygous deleterious variants in *NR6A1*. The variant spectrum includes both inherited missense as well as *de novo* loss of function, with the latter associated with a perinatal lethal phenotype characterized by bilateral renal agenesis. *In vitro* studies demonstrated partial loss-of-function for both missense variants. To investigate the role of *NR6A1* in development, CRISPR-Cas9 knockout zebrafish models targeting the orthologues *nr6a1a* and *nr6a1b* were generated. Mutants recapitulate the human phenotype, exhibiting impaired kidney development, including pronephros segmentation defects and adult kidney hypoplasia, along with axial skeletal abnormalities, cloacal anomalies, and disrupted anteroposterior expression of posterior hox genes. This study provides strong evidence linking *NR6A1* heterozygous loss-of-function variants to renal, uterine and costovertebral defects in humans.

## INTRODUCTION

The association of congenital anomalies in kidneys, uterus and the axial skeleton (vertebrae and ribs) occur more frequently than would be expected based on their respective prevalence. Unilateral renal agenesis is diagnosed in 6.5 to 11.9% of individuals with costovertebral malformations, versus 0.04% in the general population (Rai et al. 2002; Laurichesse Delmas et al. 2017). Renal and costovertebral anomalies, including scoliosis, are described respectively, in 20% (+/-9%) and 26% (+/-5%) of individuals with Mayer-Rokitansky-Küster-Hauser (MRKH) syndrome (Hall-Craggs et al. 2013; Rall et al. 2015; Kapczuk et al. 2016). Their non-random co-occurrence is also highlighted by the delineation of specific clinical entities such as VACTERL (vertebral defects (V), anorectal malformation (A), cardiac anomalies (C), tracheoesophageal fistula with or without esophageal atresia (TE), renal malformations (R) and limb malformation (L)) and MURCS (Müllerian (Mu), renal (R) and cervical somites dysplasia (CS)) in which these malformations are distinctive features. Etiologies for both VACTERL and MURCS are still mainly unknown (Jacquinet et al. 2016; Thiem et al. 2022), while reports of individuals with features of both conditions suggest the possibility of shared mechanisms for pathogenicity (Bjørsum-Meyer et al. 2016; Adam et al. 2020; Hambraeus et al. 2024). CAKUT (Congenital anomalies of kidney and urinary tract), uterine malformation and costovertebral anomalies/congenital scoliosis, in their isolated presentation and as part of VACTERL or MURCS association, are mainly sporadic. However, familial occurrence is reported in 9-15% of individuals arguing for predisposing genetic factors (Purkiss et al. 2002; Solomon et al. 2010; Rall et al. 2015; Jacquinet et al. 2016; Suman Gök et al. 2020). Variable expressivity and incomplete penetrance are observed, as illustrated in families with VACTERL or in families with renal abnormalities and/or MRKH syndrome (Hilger et al. 2012; Herlin et al. 2014), which poses challenges to the identification of new genes associated to these disorders. In isolated, or mildly syndromic CAKUT, next generation sequencing analyses have revealed high genetic heterogeneity with approximately 60 causal genes identified so far, that explain 11-20% of cases (van der Ven et al. 2018; Christians et al. 2023). In comparison, only a few genes have been involved in spondylocostal, renal and uterine malformations, including *TBX6* (deletion 16p11.2, spondylocostal dysostosis type 5 (Ma et al. 2022; Li et al. 2022)), *GREB1L* (Renal hypodysplasia type 3 #617805) (Jacquinet et al. 2020), *MNX1* (Currarino syndrome (#176470)), *SALL1, DACT1 (*Townes-Brocks syndrome 1 and 2 (#107480;#617466)) (Christians et al. 2023) and *CDX2* (Stevens et al. 2022). Unsolved familial cases suggest that new genes still have to be uncovered. By whole exome sequencing in families with recurrence of uterine and kidney anomalies, we identified *NR6A1* as a new candidate to explain predisposition to some of these various malformations.

NR6A1 (Nuclear receptor subfamily 6 group A member 1) is an orphan nuclear receptor highly conserved in vertebrates. The protein is essential for embryonic development, knockout mice being lethal at day 10.5 with severe defects in somitogenesis, abnormal chorioallantoic development, abnormal development of the ventral structures and defects in neural tube closure (Chung et al. 2001). Conditional deletion of *NR6A1* within axial progenitors in mice induces a decrease in the number of thoracolumbar vertebrae in a dose dependent manner, and modifies the timely progression of posterior hox genes expression (Chang et al. 2022). Expression data in mice and xenopus confirms its role early in development, with transient, high, widespread expression during the mid to late gastrulation (E7.5 in mice) and early organogenesis stages. Consistent with the involvement of *NR6A1* in developmental processes of the trunk, but not the tail, expression is absent in the most posterior area (posterior to the posterior growth zone in mice, near the tailbud in xenopus) and high levels were detected in the anterior and trunk regions, in the neuroectoderm and underlying mesoderm (Joos et al. 1996; David et al. 1998; Pijuan-Sala et al. 2019; Chang et al. 2022). In zebrafish, the gene is duplicated (*nr6a1a* and *nr6a1b*), and comparison of expression patterns are indicative of a conserved function for *nr6a1a* and possible neofunctionalization for *nr6a1b* (Bertrand et al. 2007). NR6A1 acts as a constitutive transcriptional repressor and was shown to inhibit pluripotency genes (e.g. *OCT4, NANOG, CRIPTO-1*) during retinoic acid mediated differentiation of embryonic stem cells (Yan and Jetten 2000; Fuhrmann et al. 2001; Gu et al. 2005; Hentschke et al. 2006). *In vitro*, NR6A1 binds as homodimers, or oligomers, with high affinity to a direct repeat of the DNA sequence AG(G/T)TCA (DR0) (Yan et al. 1997; Gurtan et al. 2013), and with lower affinity to the extended half-site TCAAG(G/T)TCA (Yan et al. 1997). The potential ligands of NR6A1 are currently unknown, hence its designation as orphan nuclear receptor.

*NR6A1* has not been associated with any human disease so far. Here, we report deleterious heterozygous variants in *NR6A1* in multiple individuals with renal, uterine, and vertebral anomalies. By inactivating the zebrafish *NR6A1* orthologs, we show that *nr6a1a/b* zebrafish mutants display kidney and axial skeletal anomalies reminiscent of those found in humans.

## METHODS

### Clinical proband samples and sequencing

Blood or salivary samples and pedigree information were collected after informed consent from individuals or their guardians. Ethical approval was obtained from the Institutional Review Boards (University of British Columbia-BC Children’s and Women’s Hospital Research Ethics Board (#H09-01228); Ethical Committee of the Faculty of Medicine of the University of Liege (#2016/133); Children’s Mercy IRB (#00000175)).

For Family 1, whole exome sequencing (WES) was performed at the University of British Columbia, Vancouver on genomic DNA isolated from saliva samples. DNA was prepared using the Ion AmpliSeq Exome target enrichment kit and sequenced on Ion Proton System sequencer, generating 150bp reads that were aligned to the human reference genome (GRCh37/UCSC hg19) for a mean of 70x average coverage. Variants were annotated and filtered using Alissa Interpret v2.1 (Agilent Technologies), to a 1% minor allele frequency in the population database gnomAD, their presence in exonic regions ±20bp of exon boundaries and a predicted change at the protein level (synonymous variants excluded). Variant in genes were prioritized by gene function (those known to be associated with CAKUT/uterine/vertebral anomalies in mammals), autosomal dominant inheritance and constraint metrics. *In silico* prediction tools such as PolyPhen-2, SIFT, PROVEAN, Mutation Assessor and Mutation Taster were used to assess the potential functional impact of missense variants and the CADD, REVEL and AlphaMissense scores (Kircher et al. 2014; Ioannidis et al. 2016; Cheng et al. 2023) were further queried for each variant of interest.

For Family 2, clinical trio exome sequencing was performed at Children’s Mercy-Kansas City on genomic DNA isolated from peripheral blood. DNA was prepared using the Kapa or TruSeq library prep and enriched using IDT xGen exome research panel supplemented with custom mitochondrial probes, and sequenced to a minimum of 7 Gb for a mean of 80x average coverage or greater on an Illumina NovaSeq 6000 (2x150 paired end reads). Bidirectional sequence was assembled, and aligned to GRCh37/UCSC hg19. Variants were filtered to a 1% minor allele frequency and prioritized by type and association using custom software, as previously described (Saunders et al. 2012; Soden et al. 2014). Briefly, candidate gene lists were generated by SSAGA and/or Phenomizer using Human Phenotype Ontology (Köhler et al. 2014) terms including Bilateral renal agenesis, Intrauterine growth retardation, Oligohydramnios, Potter facies, Pulmonary hypoplasia, and Renal agenesis with a cut off at p value of 0.5. Gene lists were imported into VIKING software to guide the analysis, however phenotype and OMIM filters were removed to look for candidates with no disease association.

For targeted sequencing, single-molecule molecular inversion probes (smMIPs; EasySeqNGS-customized Targeted Capture Kit, Nimagen) were designed to target the exons ±20 nucleotides of intronic/exonic boundaries of *NR6A1* (NM_033334.4). Except for exon 1, the entire coding sequence was covered more than x100. Single nucleotide variations were interrogated using the Seqnext (SeqPilot) software. The variants identified were confirmed by Sanger sequencing. Variants in the *NR6A1* gene are described for reference sequence NM_033334.4, which encodes for the NR6A1 reference protein NP_201591.2 using HGVS nomenclature (www.hgvs.org)

### Electrophoretic mobility shift assay

The electrophoretic mobility shift assays (EMSA) were performed essentially as described by Greschik et al. (Greschik et al. 1999) using a DR0 double stranded oligonucleotide (AGCTTCAGGTCAAGGTCAGAG) end-labbeled with 32P-dATP and Klenow enzyme. The human NR6A1 protein, as well as the R66C and M392R mutated proteins were synthesized *in vitro* using reticulocyte lysates (Promega). To that end, the coding sequences of the human wildtype NR6A1 cDNA and of the missense R66C and M392R mutants (obtained from VectorBuilder) were inserted in the expression vector pCS2-MT just downstream the SP6 promoter and fused to the MYC-tag at the C-terminus of the NR6A1 proteins. SP6 polymerase was used to produce *in vitro* the corresponding mRNA (Invitrogen mMessage Machine kit) which were next translated *in vitro* using the reticulocyte lysate following manufacturer’s instructions (Promega). Production of proteins was verified by western blot using a rabbit MYC antibody (16286-1-AP, Proteintech). For EMSA assay, proteins were incubated in buffer containing 30mM Tris-HCl (pH 7.5), 50 mM KCl, 1 mM DTT, 10 % glycerol, 1µg of poly (dI-dC) with 0.1 ng of DR0 32P-labelled probe for 20 min at room temperature. For competition and supershift assays, unlabelled oligonucleotide or MYC antibody were preincubated for 10 min with the proteins in the same buffer before addition of the DR0 probe. The reactions were loaded on a 6 % non-denaturing polyacrylamide gel in 0.5X Tris-borate-EDTA running buffer at room temperature. After the run, gels were dried and autoradiographed.

### Generation of Crispr-Cas9 *nr6a1a/b* zebrafish mutants

All studies performed in Zebrafish were approved by the ethical committee of Liège university (protocol 2404).

CRIPSR-Cas9 mediated genome editing was used following the IDT protocol ‘Zebrafish embryo microinjection Ribonucleoprotein delivery using the AltR™ CRISPR-Cas9 System’, contributed by Jeffrey Essner, PhD ([CSL STYLE ERROR: reference with no printed form.]). CRISPR-Cas9 crRNA targeting exon 3 of *nr6a1a* (GRCz11Ensembl Transcript ID: ENSDART00000191653.1; exon 3, 5’ - CAGGACTGCACTATGGCATT -3’) and the crRNA targeting exon 3 of *nr6a1b* (GRCz11Ensembl Transcript ID: ENSDART00000011096.8; exon 3 5’-GCACTATGGTATTATTTCCT-3’) were designed using ChopChop to generate F0 mosaic embryos. The Alt-R-crRNA, Alt-R CRISPR-Cas9 tracrRNA and Alt-R Cas9 Nuclease V3 were obtained from Integrated DNA technologies.

The null mutation nr6a1aulg083 is a 5bp deletion in exon 3 (ACCGCGCCACAGGACTGCACTATGG) leading to a frameshift in the coding sequence and generating a restriction enzymatic Btslv2 site. For genotyping, the 249pb fragment was amplified using the primers CAGGCTGAGCAGCGCTCTTGTC and TCCAGCGTGTGTGTTTCTCTCAC and terra Taq polymerase (Takara) followed by Btslv2 digestion. The null mutation nr6a1bulg085 is an indel (CCTGCACTATGGTATTATTTCCTG to CCTGCATGCTATATATTTCCTG) leading to a frameshift and disrupting a BsgI restriction site. Genotyping this variant was done by amplifying the 387 bp fragment using primers TGCCCAATGGTTGGTTTGGCTACA and GTGTGTCTCACCCTTGCGGTTCATG followed by BsgI digestion.

### Dissection and measurement of the adult zebrafish kidney after labeling of the proximal tubule with fluorescent Rhodamine-dextran

15 months-old adult fish were anesthetized in tricaine and injected with 10 µl of Rhodamine-dextran 3.5% in the peritoneal cavity. Three days later, the kidney was dissected as described previously (McCampbell et al. 2014). The adult kidney (mesonephros) was gently detached from the dorsal wall, placed on a glass slide, flattened and covered by a glass coverslip. Rhodamine-dextran compounds, incorporated by the epithelial cells in the proximal comvoluted tubule (PCT), allow: - 1) to better delineate the mesonephros colored in dark-red; - 2) to functionally assess the endocytic uptake by the proximal tubule cells. Pictures were taken with Leica fluorescent binocular (M165). Area measurements were performed with Fiji (ImageJ). For each fish, the ratio of kidney surface (μm2) to fish length (mm) was calculated and then normalized to that of wildtype fish. Graph and statistics (parametric t-test and p-value) were obtained with JMP-Pro 17.2.

### MicroCT scan

Adult fish were anesthetized in tricaine and then incubated in 4 % paraformaldehyde for 2 days at 4°C. Fish were rinsed three times in PBS prior to being imaged for skeletal structures analysis. Imaging was done on a Bruker Micro-CT Skyscan 1172/G. The fish were fixed in home-made adapted cylindrical EPS containers and PBS was added to avoid dehydration over the duration of the scan. After mounting the sealed container on the scanner’s sample stage, a relaxation time of 45 minutes was left prior to scanning. The voltage and intensity of the X-ray source were set at 100 kV and 100 µA respectively and a 0.5 mm Al filter was used. The exposure time was set at 300 ms and acquisitions proceeded over 360° with steps of 0.4°. The detector is a SHT 11Mp camera, set with 4x4 binning to obtain projections of 1000 by 666 pixels in size.

After this microCT scan, the same fish were stained by incubation in Lugol 2.5 % for one day at room temperature, washed three times in PBS for 5 minutes and scanned again for analysis of soft tissues. The same conditions were used for the setup and the scans except that the rotation step was reduced to 0.2° and the imaging limited to the abdominal zone.

After scanning, reconstruction was performed on the Bruker software NRecon. Further image analyses were performed upon processing the data with the visualization software Avizo with custom-made computing modules.

### Whole-Mount Zebrafish in situ hybridization

Whole-mount is situ hybridization (WISH) of zebrafish embryos using digoxigenin- and FITC-labelled probes were performed as described previously (Hauptmann and Gerster 1994; Thisse and Thisse 2008). The expression patterns of *trpm7, gata3, clcnk, slc12a3, xirp2a* and *tbx5* were previously reported (Mercader et al. 2006; Wingert et al. 2007; Chen and Galloway 2014). The 3’ part of cDNA of *hoxa9b* (GTGGTCCAGCAGCAGTCTCGTG and GGATCTAGCTTCGTCTCCGCAGG), *hoxd11a* (CGCAAGTCCAACCTGTTCGC and GGGCTCGAGTGCGACAAAGTC), *hoxb13a* (GTGAGCGTGCTATGACCACC and GGAGACTATCGTGTCGCGG) and *evx1* (AGCTTTGGGCACTTTGGCAGT and GCTGCACATTTCGGTGTGCTGC) were amplified by PCR using the corresponding primers and the cDNA were inserted downstream the T3 promoter in the reverse orientation for the production of antisense labelled RNA by *in vitro* transcription. WISH were performed on clutches of 40 or 80 embryos respectively obtained from incross of *nr6a1a^+/-^* fish or of *nr6a1a^+/-^nr6a1b^+/-^* fish or of *nr6a1a^+/-^nr6a1b^-/-^* fish. After WISH, about 20 stained embryos were photographed and genotyped as described above. Graph and statistics (student’s t-test and p-value) were obtained with JMP-Pro 17.2.

## RESULTS

### Clinical presentation

In Family 1, the proband (individual II:1) presented with Mayer-Rokitansky-Küster-Hauser syndrome (Fig. 1a and Table 1), along with a single pelvic kidney, ectopic ovaries and a history of vesicoureteral reflux requiring surgery. Endocrinological workup was normal with no hyperandrogenism. Spine and chest x-rays revealed the presence of 11 thoracic vertebrae instead of 12, with only 10 ribs on the right, and severe hypoplasia of the eleventh rib on the left (Fig. 1b). Standard karyotype was 46,XX and microarray (180k SNP array) was negative. The family history was significant for a sister (individual II:2) who was diagnosed with right renal hypoplasia and left compensatory renal hypertrophy. She had no uterine malformation. In addition, the mother (individual I:2) had a hemi-uterus with a single median pelvic kidney and one single right ovary. She underwent kidney transplantation in her 50s due to end-stage renal insufficiency. On imagery, the native kidney was atrophic with loss of differentiation.

**Fig. 1.**
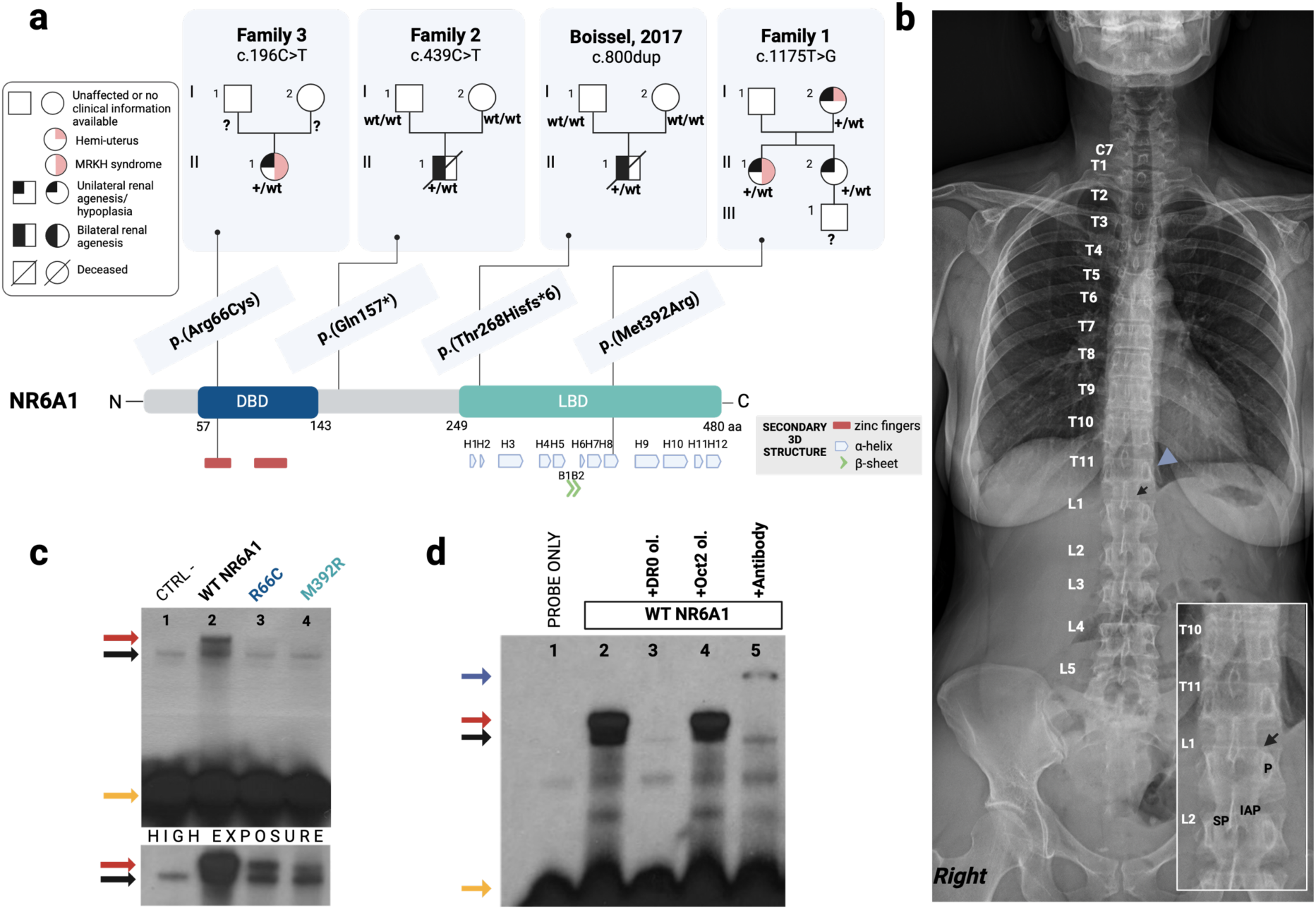
Heterozygous variants in *NR6A1* in four families with renal agenesis/hypoplasia with/without uterine and skeletal anomalies. a. Pedigree and localization of the truncating and missense variants along the NR6A1 protein (NM_033334.4). The genotype of relatives is mentioned as “?” if not tested. NR6A1 is depicted with its main functional domains and 3D structure. The DNA-binding domain (DBD in blue) include two zinc fingers (red rectangles). The 3D structure of the ligand binding domain (LBD in turquoise) is composed of twelve alpha-helices (H1-H12) and two beta-turns (B1-B2). b. X-ray of the spinal column (Family 1; individuals II:1) showing 11 thoracic vertebrae (T), 10 ribs on the right and a 11th hypoplasic rib on the left (blue arrow head), and a normal number of five lumbar vertebrae (L). The black arrows point to the interspace of the facet joint visible between the last thoracic vertebrae and the first lumbar vertebrae given its orientation. IAP=inferior articular process, P= pedicle, SP= spinous process c. Electrophoretic mobility shift assay with ^32^P-radiolabeled DR0 consensus sequence, showing lower complex formation for the mutated hsNR6A1 proteins compared to controls (red arrow). An additional band (black arrow) is detected in the four samples corresponding to non-specific interaction of DR0 probe with proteins of reticulocyte lysate. The yellow arrow points to the unbound labeled DR0 oligonucleotides. The negative control (CTRL-) corresponds to incubation of probe with reticulocyte lysate without any mRNA. WT NR6A1, R66C and M392R correspond to wildtype and mutant NR6A1 proteins synthesized in reticulocyte lysates. d. Competition analyses and supershift assay. The red arrow points to the band formed by the DR0-NR6A1 complex, the black arrow points to non-specific protein-DNA complexes from the reticulocyte lysate, the yellow arrow points to the unbound labeled DR0 oligonucleotides, and the blue arrow points to the shifted complex in presence of anti–Myc antibody. Created with BioRender.com

**Table 1.** Phenotype and genotype of individuals identified with a heterozygous variant in *NR6A1*.

|  | Renal phenotype | Uterine phenotype | Other phenotypes | Nucleotide change | Protein change | MAF gnomAD<sup>1</sup> | CADD score | REVEL score | Alphamissense |
| --- | --- | --- | --- | --- | --- | --- | --- | --- | --- |
| **Family 1** |  |  |  | c.1175T>G | p.(Met392Arg) | NR | 29.6 | 0.97 | 0.996 |
| **I :2** | URA, ectopic kidney | Hemi-uterus | Single right ovary |  |  |  |  | (deleterious strong) | (deleterious strong) |
| **II :1** | URA, ectopic kidney, VUR | Uterine agenesis | 11 TV, 10 ribs on the right, hypoplasia of the left 11<sup>th</sup> rib; ectopic ovaries |  |  |  |  |  |  |
| **II:2** | Right renal hypoplasia | Normal | - |  |  |  |  |  |  |
| **Family 2** |  |  |  |  |  |  |  |  |  |
| **II :1** | BRA | - | - | c.439C>T | p.(Gln147*) | NR | 47 | - | - |
| **Family 3** |  |  |  |  |  |  |  |  |  |
| **II :1** | URA | Uterine agenesis | - | c.196C>T | p.(Arg66Cys) | 8.674×10<sup>-6</sup> | 33 | 0.92 (deleterious moderate) | 0.992 (deleterious strong) |

URA=unilateral renal agenesis, BRA= bilateral renal agenesis, TV= thoracic vertebrae, VUR= vesicoureteral reflux <sup>1</sup>gnomad v4.1

Submission to Genematcher (Sobreira et al. 2015) allowed to identify the second case (individual II:1 in Family 2), a male newborn who died very shortly after birth due to respiratory insufficiency in the context of bilateral renal agenesis, anhydramnios and intrauterine growth retardation, which were identified at the second trimester ultrasound. X-rays and autopsy were not performed. Family history was unremarkable.

Finally, individual II:1 in Family 3 presented with MRKH syndrome, a left ectopic kidney, and a history of vesicoureteral reflux requiring surgery. Spine X-ray was normal. Karyotype was 46,XX.

### Identification of *NR6A1* heterozygous variants

Exome sequencing was performed on the three affected members of Family 1. Rare heterozygous variants shared by all three relatives were filtered and prioritized. The missense variant c.1175T>G; p.(Met392Arg) in *NR6A1* was identified as the most compelling candidate given the crucial role of *NR6A1* in embryonic development, alongside phenotypic resemblances in the proband to the vertebral anomalies observed in conditional knockout (Chang et al. 2022). The variant c.1175T>G is not observed in gnomAD v4.1 and is predicted to be deleterious by several *in silico* tools (Table 1). It affects the ligand binding domain (LBD) of NR6A1, substituting a highly conserved methionine that is hydrophobic and uncharged by a hydrophilic and charged residue, an arginine. The Grantham’s distance between both aminoacids is 91 [0-215]). The hydrophobic residue, conserved amongst nuclear receptors, contributes to the folding of the protein by forming van der Waals contacts between the 8^th^ and 10^th^ α-helices (Wurtz et al. 1996; Greschik et al. 1999). Hence, changes of physicochemical properties of this amino acid likely alters the tertiary structure of the protein which may impact the affinity of the DNA binding, the homodimerization or interaction with co-repressors.

Trio-WES in Family 2 identified a *de novo* heterozygous variant, c.439C>T; p.(Gln147*), in *NR6A1* in individual II:1 (Fig. 1a). Interestingly, another *de novo* heterozygous truncating variant, c.800dup;p.(Thr268Hisfs*6) was previously reported by Boissel et al. (Boissel et al. 2017) in a male fetus with bilateral renal agenesis, absence of the ureters, a unique umbilical artery and a retroesophageal right subclavian artery. Both variants generate premature termination codons, and hence, are likely to induce nonsense-mediated mRNA decay. Structural analysis suggests that any residual mRNA would encode a protein missing the entirety or most of the LBD domain, resulting in loss-of-function of the protein (Okumura et al. 2013). *NR6A1* is highly constrained for loss-of-function variation in gnomAD, with observed/expected value (LOEUF:0.19) (Chen et al. 2024), indicating such variants are not tolerated in the population.

To identify additional individuals with *NR6A1*-associated disease, we performed targeted sequencing of *NR6A1* in a cohort of 68 individuals with MRKH and identified a heterozygous variant, c.196C>T; p.(Arg66Cys) in individual II:1 of Family 3 (Fig. 1a). It is unknown if the variant was *de novo* or inherited as parental samples were unavailable for segregation studies. The c.196C>T; p.(Arg66Cys) variant is rare in the population database gnomAD (11 heterozygous males/801566 and 3 heterozygous females/812472) and is predicted deleterious *in silico* (Table 1). The variant is located in the DNA binding domain, in the first zinc finger subdomain, and affects a highly conserved arginine, that is substituted by a cysteine (Grantham’s distance=180 [0-215]), possibly altering the interaction with the DNA.

### Decreased DNA binding affinity of Arg66Cys and Met392Arg NR6A1 proteins *in vitro*

In order to ascertain whether the two missense variants we identified could alter the binding of NR6A1 to its target DNA and therefore prevents its transcriptional activity, we performed an electrophoretic mobility shift assay (EMSA). The native or the mutated form of the hsNR6A1 protein fused to a Myc-tag polypeptide were *in-vitro* translated and their concentration were assessed by western blot (Supplementary Fig. 1). As probe we used a radiolabeled oligonucleotide consisting of a direct repeat with zero spacing (DR0) of the consensus sequence AGGTCA. The probe was incubated with either the native hsNR6A1 or Arg66Cys and Met392Arg hsNR6A1 proteins (Fig. 1c). Whereas a strong complex was formed on the DR0 probe with the native NR6A1 (lane 2), the complexes formed with the mutated NR6A1 proteins Arg66Cys (lane 3) and Met392Arg (lane 4) were almost undetectable, only a weaker signal was visible upon extended exposure. The DNA binding specificity was verified by a competition assay (Fig. 1d); indeed the addition of a 125 fold excess of unlabeled DR0 blocked formation of the protein-probe complex (lane 3) while the addition of an unlabeled Oct2A-binding oligonucleotide had no effect (lane 4). The presence of NR6A1 protein in the complex was further confirmed with the observation of a supershift in presence of anti–Myc antibody (lane 5). Together these results showed that while native NR6A1 binds specifically DR0 sequence with high affinity, the mutations Arg66Cys and Met392Arg strongly decrease this affinity. It therefore suggests that the identified missense variants can prevent NR6A1 activity.

### Loss of Nr6a1a/b in zebrafish cause skeleton anomalies and abnormal kidney morphology

To evaluate the impact of NR6A1 loss-of-function, in particular in urogenital tract and skeleton formation, we inactivated the zebrafish orthologs *nr6a1a* and *nr6a1b* by CRISPR-Cas9. The higher degree of sequence homology between *nr6a1a* and *NR6A1* and the similarities in the pattern of *nr6a1a* expression compared to other species suggested that most of the conserved functions would be controlled by *nr6a1a*. However, the high similarity in the DNA binding domain between Nr6a1a and Nr6a1b did not exclude overlapping function in gene regulation. Moreover, although expression of *nr6a1b* occurs later, after 10 hours post fertilization (hpf), and is more restricted (Supplementary Fig. 2), both genes are expressed in same regions of the trunk (neural tube, paraxial and lateral mesoderm) during somitogenesis. In order to detect compensatory effect of the duplicated genes, phenotypes of single and double null mutants for *nr6a1a* and *nr6a1b* were analyzed.

No alteration was noted in *nr6a1b* ^ulg085-/-^ embryos and these mutants reached adulthood without any obvious defects. However, only a low percentage (around 4-10%) of *nr6a1a^-/-^* survived until adulthood. *Nr6a1a ^ulg083-/-^* homozygous embryos displayed transient pericardial oedema from 30 to 56 hpf. The homozygous mutants that reached adulthood displayed visible morphological anomalies such as shorter trunk and absence of the anal fin (Fig. 2a). Unilateral or bilateral aplasia or hypoplasia of the pectoral fin were also observed in a low percentage (6%) of *nr6a1a^-/-^* larvae. Homozygous mutants were slightly shorter compared to siblings (9.2 % shorter in average), with a disproportionate reduction of the anterior region of the trunk (10% reduction in average relative to the size) (Fig. 2a). The homozygous females were infertile. Double *nr6a1a ^ulg083^/b ^ulg085^* homozygotes died before 16 days post fertilization (dpf). They presented the same morphological anomalies than the *nr6a1a^-/-^* mutants with an increasing penetrance of the pectoral fin phenotype supporting a compensatory role for *nr6a1b* in some tissues (Supplementary Fig. 3).

**Fig. 2.**
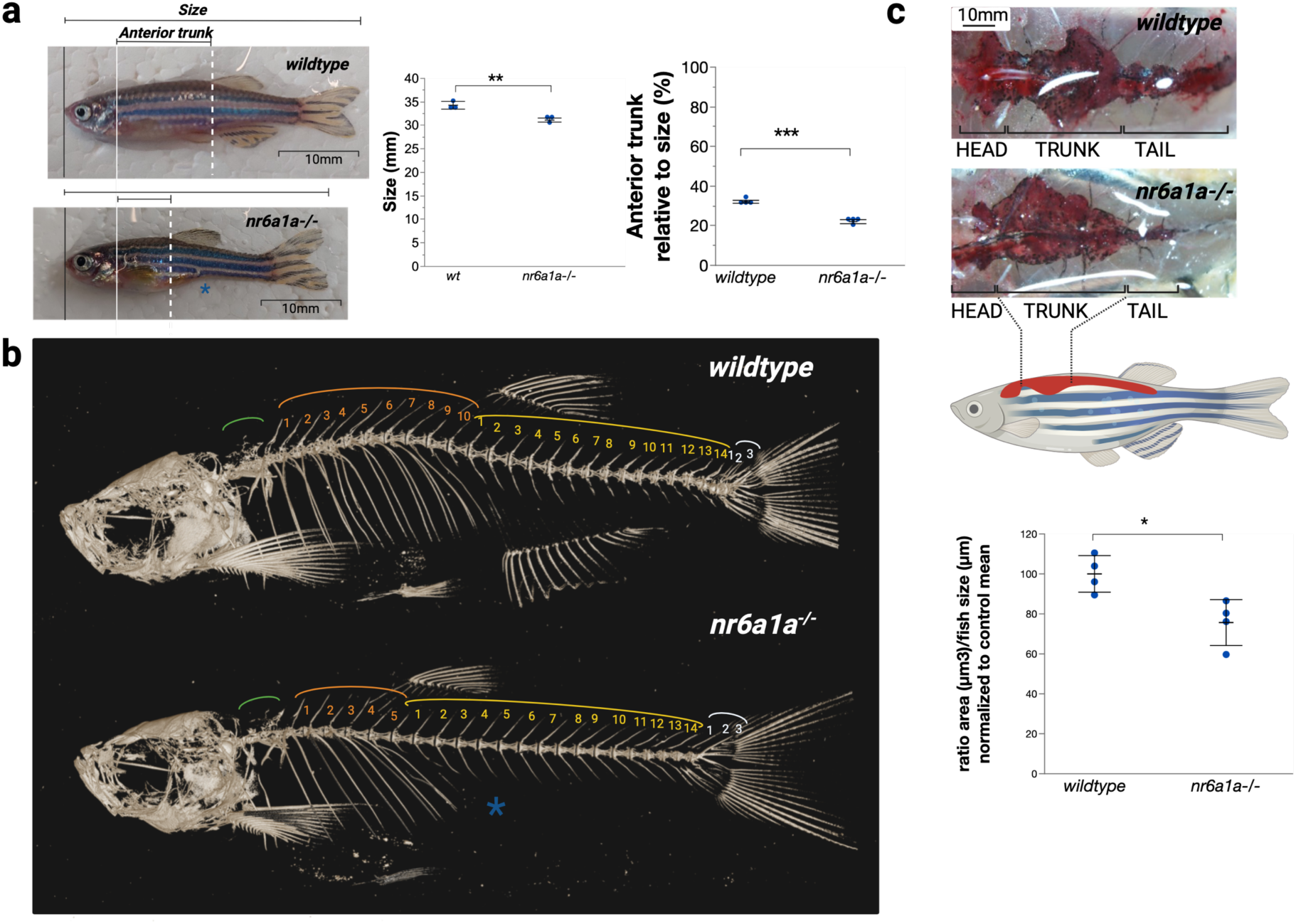
Smaller size, decreased number of ribs-bearing vertebrae and abnormal size and morphology of the kidney in *nr6a1a^-/-^* adult zebrafish. a. Comparison of the size of wildtype (n=3) and mutant (n=3) siblings at 5 month ½ (mean size wildtype: 34.295mm; mean size mutants:31.125mm: difference: 3,17mm (CI95%:1.47-4.87) (p value=0.0095)). Comparison of the length of the anterior part of the trunk relative to the fish size between wildtypes (n=4) and mutants (n=4) (mean ratio wildtype 32.048 mm; mutants:22.036 mm; difference 10.01 mm (CI95%:8.43-11.59) (p value <0.0001). b. X-ray microtomography of 3-month-old wildtype and mutants. The number of rib-bearing precaudal vertebrae (orange) is decreased in mutants (#5 instead of 10 in wildtypes). The rib-bearing precaudal vertebrae are recognizable by the presence of ribs, and the absence of hemal arches and spines, that are them specific of the caudal vertebrae (Bird and Mabee 2003). The number of caudal vertebrae (yellow, and light blue for the caudal fin vertebrae) is normal in mutants (#17). Absence of the anal fin (beginning at the 1st caudal vertebrae in wildtype) in mutants (*). The weberian vertebrae (vertebrae 1 to 4) are highlighted in green and seems normal although not investigated in details. c. Mesonephros morphology of 15 month-old wildtypes and mutants. The mutants present an abnormal shape of the mesonephros, dissected after injection of Rhodamine-dextran for visualization. Compared to wildtypes (n=4), the surface of the mesonephros relative to the fish size (after normalization by the control mean), is decreased by 24.35% (CI95%:6.15-42.54) (p value=0.017) in *nr6a1a^-/-^*mutants (n=4). Created with BioRender.com

To characterize the skeletal defects, MicroCT scans were then performed on adult *nr6a1a*^-/-^ mutants and wildtype females. *Nr6a1a*^-/-^ mutants presented a reduced total number of vertebrae from 31 in wildtypes to 26 (+/-1) in mutants (n=4). Interestingly, the five missing vertebrae are located in the trunk and correspond to precaudal vertebrae from which the ribs originate from. This is consistent with the shortened trunk region we observed (Fig. 2b and Supplementary Fig. 4). The dorsal fin radials start at the level of the penultimate precaudal vertebrae, corresponding to the 4^th^ and 9^th^ non-Weberian precaudal vertebra in mutants and wildtypes, respectively. The axial skeleton of some mutants presents other defects, such as abnormal, irregular shape of the ribs and unfused hemal arches of some caudal vertebrae (Supplementary Fig. 4). Moreover, we confirmed the absence of the anal fin rays (Fig. 2b).

Because of anomalies of the female genital tract in humans with *NR6A1* variants and infertility in *nr6a1a^-/-^* zebrafish females, we questioned the possibility of gonadal duct anomalies in surviving adults. Formation of the gonadal duct starts around 25dpf in teleost, with firstly the formation of the upper part of the oviduct in continuity with the ovarian cavity, progressive elongation and secondarily, junction to a lower part resulting from invagination and cavitation of the urinogenital papillae around 50dpf (Suzuki and Shibata 2004; Kossack et al. 2019). Incubation of whole fish in a lugol solution allow for visualization of soft tissue by MicroCT scan. Using this contrast agent, we were able to visualize the oviducts. The oviduct seemed formed in the *nr6a1a^-/-^* females, however the channel looked obstructed compared to controls (Supplementary Fig. 5). Unfortunately, the precocious lethality of the double mutant did not allow to address the impact of a total loss-of-function of *nr6a1* alleles on the gonadal tract.

As congenital anomalies of the kidneys were present in humans (aplasia/hypoplasia/ectopia), we investigated the kidney structure in the adult *nr6a1a^-/-^* mutants. To easily visualize the renal tissue, we injected intraperitoneally a solution of Rhodamine-dextran in 4 wildtypes and 4 *nr6a1a^-/-^* mutants. This small molecule is uptaken by the kidney proximal tubules within a day post injection. It colors the organ in red, and allows a functional assessment of the proximal tubules (McCampbell et al. 2014). The adult zebrafish kidney is a flat organ located on the dorsal body wall. When viewed from a ventral perspective, the wildtype kidney has a distinctive curved morphology, consisting of head, trunk and tail regions. This typical shape is perturbed in the *nr6a1a^-/-^* mutants, the trunk segment being larger. To determine if there was a change in the size of the kidney relative to the fish size, we measured the overall surface of the kidney and made a ratio on the length of the fish. Hence, we showed a decrease by 24% of the kidney size in the *nr6a1a^-/-^* mutants compared to wildtypes (p-value=0.017) (Fig. 2c). The proximal tubule of the *nr6a1a^-/-^* mutants seemed functional as we could observe uptake of Rhodamine-dextran as in wildtypes (Supplementary Fig. 6).

Together, our data show that total inactivation of NR6A1 orthologs is embryonically lethal and that inactivation of Nr6a1a alone is sufficient to induce costovertebral defects and kidney hypoplasia. Those anomalies being similar to those observed in human individuals with *NR6A1* variants, it strongly suggests that these variants are responsible for their syndrome.

### Nr6a1a/b are required for proper pronephros development

Given the developmental origin of the malformations in individuals with *NR6A1* variants and as we showed that *NR6A1* is essential for embryonic development, we further investigated the function of the gene in kidney development. The zebrafish pronephros, as the earliest nephric stage, contains two nephrons sharing numerous genetic, structural, and functional aspects with the mammalian nephrons. From 24hpf, it consists of glomeruli and of tubular epithelium extending from the glomerulus to the cloaca, where the gastrointestinal tract also opens up. Interestingly, the terminal end of the pronephros (named pronephric duct, PD) was abnormal in *nr6a1* mutants with variable expressivity as shown by brightlight microcoscopy at 5 dpf (Fig. 3a). The double homozygous mutants were the most severely affected, with some larvae showing a blind end of the digestive tract and an indistinguishable pronephric duct orifice. A milder phenotype was observed in the simple *nr6a1a^-/-^* mutants, with presence of the gut opening but abnormal cloaca with a not well-defined end of the pronephric duct.

**Fig. 3.**
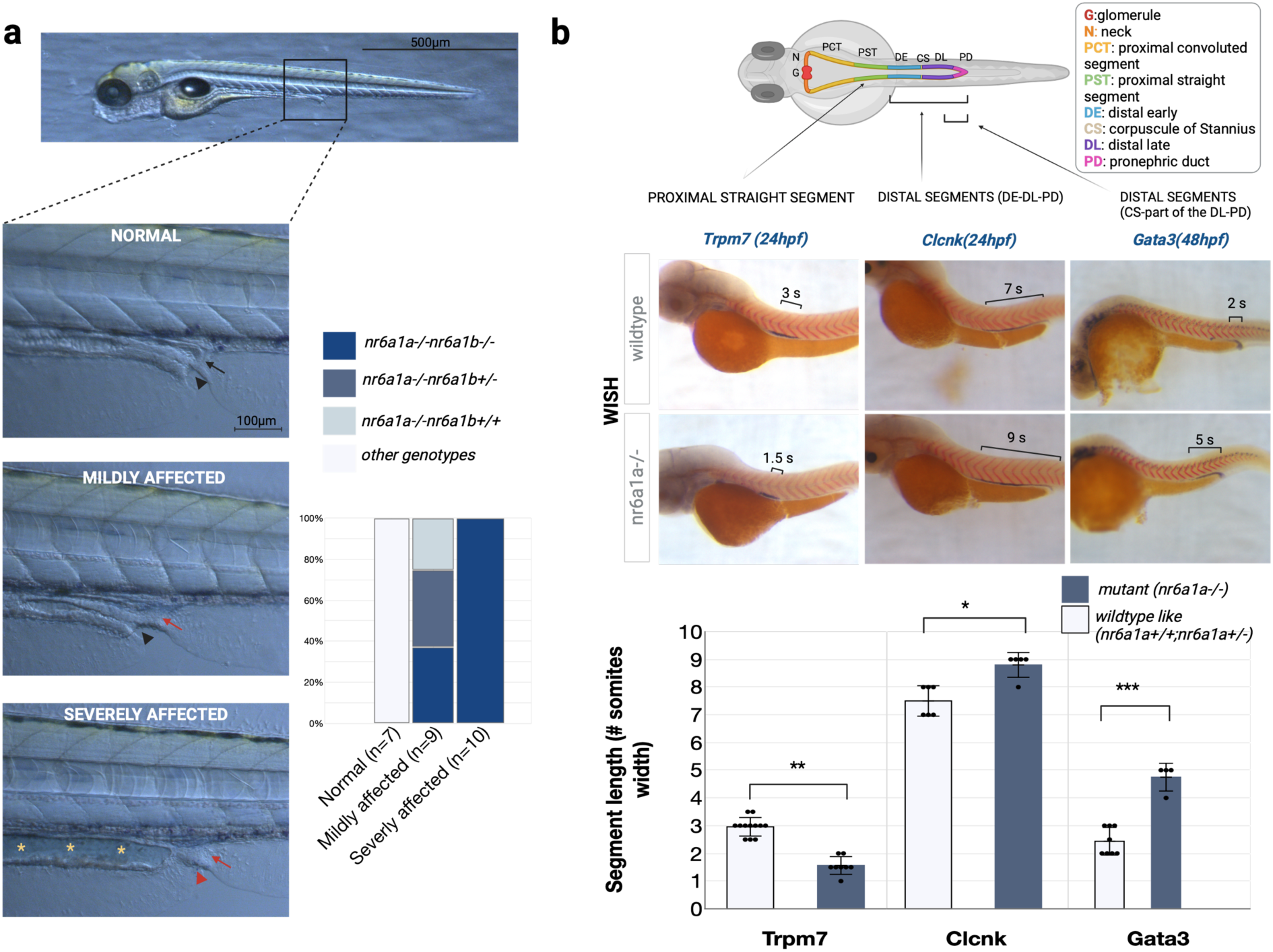
Defects in cloacal morphogenesis and pronephros patterning in *nr6a1a^-/-^* and *nr6a1a^-/-^ nr6a1b^-/-^* mutants. a. Brightfield images of the cloaca at 120hpf in embryos of *nr6a1a^+/-^nr6a1b^+/-^* incross. Classification of the phenotype as normal if the cloaca presents a typical inverted U-shaped with a visible pronephric duct in its terminal part (black arrow), and a visible opening of the gut (black arrow head); mildly affected if the lumen of the gut reach the cloaca (black arrow head) whereas the terminal part of the pronephric duct is not visible (red arrow); severely affected if both ends are not visible (red arrow and red arrow head). Some severely affected embryos present dilation of the gastrointestinal tract (yellow stars), suggesting imperforation. 26 embryos were selected for genotyping (normal (n=7); mildly affected (n=9); severely affected (n=10)) Embryos showing normal morphology are wildtype or heterozygous for *nr6a1a* and show variable *nr6a1b* genotypes. Affected embryos are all *nr6a1a^-/-^* with severely affected embryo being homozygous for both *nr6a1a* and *nr6a1b*. b. Double whole-mount in situ hybridization with the somite boundary marker *xirp2a* and the pronephros segment markers *trpm7, clcnk* and *gata3* in embryos from *nr6a1a^+/-^* incross. The width of one somite is used as reference to measure the length of the expression domain. Expression in *nr6a1a^-/-^*mutants compared to wildtype/heterozygotes is increased with a difference of +2.3 somites (p<0.0001) for *gata3* (n=4 for *nr6a1a^-/-^* and n=8 for *nr6a1a^+/-or^ ^+/+^*),+1.3 somites (p=0.0022) for *clcnk* (n=5 for *nr6a1a^-/-^* and n=6 for *nr6a1a^+/-or^ ^+/+^*), and -1.4 somites (p<0.0001) for *trpm7* (n=8 for *nr6a1a^-/-^* and n=12 for *nr6a1a^+/-or^ ^+/+^*). Created with BioRender.com

From 24hpf onwards, the tubular epithelium of the zebrafish pronephros is subdivided into segments that are functionally homologous to segments of the mammalian nephron: two proximal tubule segments (proximal convoluted tubule (PCT) and proximal straight tubule (PST)), connected to the glomerule (G) by the neck segment (N) and two distal tubule segments (distal early (DE) and distal late (DL)) connected to the cloaca by a short segment (the pronephric duct (PD)) (Fig. 3b) (Wingert et al. 2007). Double whole-mount in situ hybridization (WISH) with segment-specific marker gene and the somite boundary marker *xirp2a* (Wingert et al. 2007) were performed at 24hpf and/or 48hpf on embryo from single and double heterozygous mutants incross. *Clcnk* and *gata3* are normally expressed in the distal segments (*clcnk*: DE-DL-PD; *gata3:* distal part of the DL-PD and corpuscule of Stannius (CS)). The expression of both markers is extended anteriorly by around 2 somites width in *nr6a1a^-/-^*(Fig. 3b and Supplementary Fig. 7). Hence, *clnck* expression that usually starts at the level of the 9^th^ somites is seen as anterior as the 7 somites in the mutant (average extension of 1.3 somites width (CI 95%:0.62-1.98). *Gata3* is extended anteriorly by 2.3 somites width in average (CI 95%:1.57-3.06). Conversely, expression of *trpm7*, which labels the proximal straight segment, is reduced. *Trpm7* is expressed from the sixth somites, it extends to the 9^th^ somite in wildtype but stops between the 7^th^ and the 8^th^ somites in homozygous mutants (average length decreased by 1.4 somites width, CI95%:1.08-1.71). Finally, an extended expression was observed in mutants (selected as showing no *tbx5* expression) compared to wildtypes for *evx1*, a gene normally expressed at 24hpf in the posterior gut, the cloaca and the lateral mesoderm surrounding the end of the pronephric duct (Supplementary Fig. 7) (Thaëron et al. 2000). No differences were observed in the expression of markers labeling the glomerules (*wt1a*, *nephrin*) or PCT (*slc20a1*) in *nr6a1a^-/-^* mutants compared to wildtype. Double homozygous (*nr6a1a^-/-^*; *nr6a1b^-/-^*) have similar expression of *gata3* to the simple *nr6a1a^-/-^* mutant in standard conditions (Supplementary Fig. 7). These results show the involvement of *nr6a1a* in nephron patterning with a substantial effect on the intermediate and distal segments.

### Nr6a1a/b loss-of-function modify posterior Hox genes expression in zebrafish

The phenotype observed in the patterning of the pronephros as well as the missing vertebrae are reminiscent of antero-posterior patterning defects. As in vertebrates Hox genes provide the major positional information along the anteroposterior axis, we investigated hox genes patterning in *nr6a1a/b* mutants. We focused on *Hoxa9–13* and *Hoxd9–13* as these posterior Hox determines notably the patterning of the female genital tract (Taylor et al. 1997; Raines et al. 2013), as well as vertebrae and ribs patterning (Hubert and Wellik 2023), metanephros formation and nephron segment identity (Wellik et al. 2002; Drake et al. 2018) in mammals. Posterior hox paralogs demonstrate collinear expression in the trunk along the anteroposterior axis of the zebrafish embryo; they are expressed in ventral and lateral mesoderm with anterior boundaries depending on each particular hox gene (van der Hoeven et al. 1996; Sordino et al. 1996). WISH were first performed for *hoxa9b* in 31 hpf embryo from *nr6a1a^+/-^nr6a1b^+/-^*incross. Compared to wildtypes, the domain of expression of *hoxa9b* was extended anteriorly when *nr6a1a* was inactivated (*nr6a1a^-/-^).* The loss of one or two *nr6a1b* allele did not exacerbate the phenotype, showing that *nr6a1a* is the main contributor to the phenotype at least for *hoxa9b* (Fig. 4a). Similar results were shown for *hoxa10b* at 18hpf (18 somites stage), with no differences following *nr6a1b* inactivation alone (Supplementary Fig. 8). As *nr6a1b* could still compensates *nr6a1a* for other targets, we systematically investigate the expression of other posterior hox gene in a *nr6a1b^-/-^* genetic background. WISH were performed at 18hpf and 24 hpf for *hoxa10b (n=18), hoxa11b (n=17), hoxa13b (n=17) or hoxd11a (n=17).* Similar to *hoxa9b*, an anterior shift of expression was observed for those markers. Embryos were genotyped based on their phenotype and we showed that 100% of the embryo with a pattern of expression shifted anteriorly were homozygous for *nr6a1a* and that all the *nr6a1a -/-* embryo exhibit this shift (Fig. 4b and Supplementary Fig. 8). Interestingly, *hoxb13a* whose expression is restricted to the posterior extremity of tail in wildtype embryos, is ectopically expressed in the double *nr6a1a;nr6a1b* mutants at 24hpf (Fig. 4c). Indeed, ectopic expression appears just above the yolk extension in the posterior endoderm and/or mesoderm with the inactivation of both *nr6a1a* and *nr6a1b*. Very mild ectopic expression was detected in *nr6a1a* single mutants (Fig. 4c). Finally, we tested several paralogues of the anterior and central hox clusters *(hoxb1a, hoxa2b, hoxb3a, hoxd3a, hoxb4a* at 21 hpf*; hoxb6b, hoxb8b* at 31hpf) and we did not detect any differences between mutants and wildtypes. These findings indicate that *nr6a1a* globally regulates the expression pattern of several posterior *hox* genes from paralogous groups a,b and d in zebrafish, resulting in ectopic expressions that extend anteriorly in mutants.

**Fig. 4.**
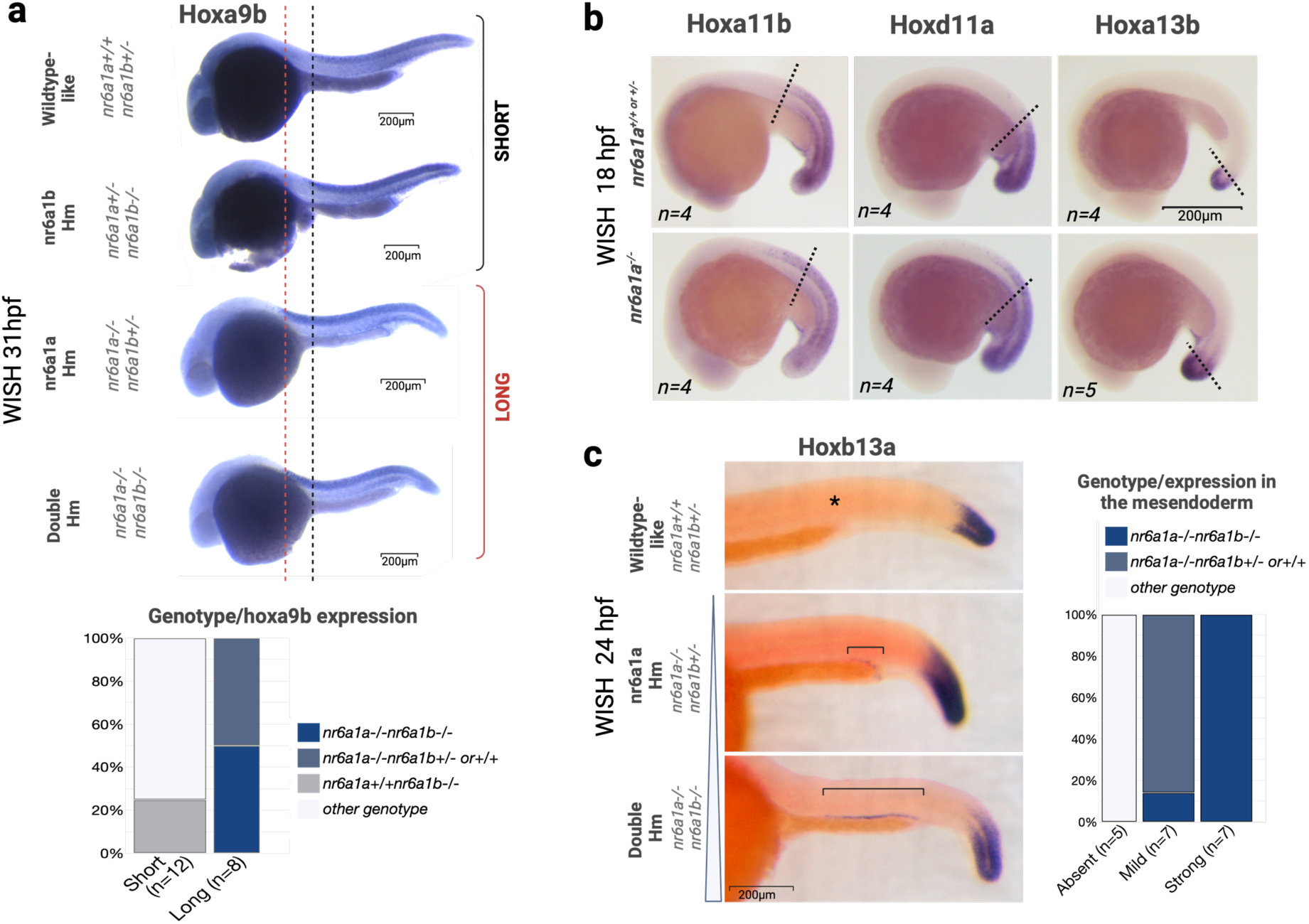
Abnormal patterning of posterior Hox in *nr6a1a^-/-^* and *nr6a1a^-/-^nr6a1b^-/-^* mutants. a. Whole-mount is situ hybridization with *hoxa9b* at 31hpf in embryos from an incross between *nr6a1a^+/-^ nr6a1b^+/-^* and *nr6a1a^+/-^nr6a1b^-/-^* fish. A longer anterior axial staining (neural tube) was observed in ¼ of the stained embryo (18/68). Genotyping performed in 20 embryos (short phenotype: n=12; long phenotype: n=8). b. Whole-mount in situ hybridization with *hoxa11b, hoxd11a, hoxa13b* at 18hpf in embryos from an incross between *nr6a1a^+/^-nr6a1b^-/-^* showing an extended anterior staining in *nr6a1a^-/-^* mutants. c. Whole-mount is situ hybridization with *hoxa13b* at 24hpf in embryos from an incross between *nr6a1a^+/-^ nr6a1b^+/-^* double heterozygous. Classification of the embryo following the extend of the staining in the posterior mesendoderm (from absent to severely extended). Genotyping was performed for 19 embryos. Created with BioRender.com

## DISCUSSION

In this study, we report evidence that strengthens the association between the *NR6A1* gene and congenital anomalies in humans. Our findings shed light on the role of *NR6A1* in human development, heterozygous deleterious variants in the gene predisposing to renal agenesis/hypoplasia, spondylocostal and uterine malformations. The alteration in kidney and skeleton observed in our knockout *nr6a1a/b* zebrafish model support the gene’s causality. Functional assessment of the two *NR6A1* missense variants identified in human subjects revealed decreased binding to the consensus DNA sequence, confirming a partial loss-of-function for the mutated proteins. Meanwhile, the two protein-truncating variants, found in fetuses, likely result in mRNA decay and *NR6A1* haploinsufficiency, or alternatively would produce a truncated protein lacking the LBD domain. Individuals harboring heterozygous protein-truncating variants exhibit severe defects like bilateral renal agenesis, while missense variants are linked to non-lethal malformations such as unilateral renal agenesis, renal hypoplasia, and kidney ectopia with or without uterine and skeletal anomalies. In our zebrafish model, we observed a dose-sensitive effect, where some phenotypes become progressively more severe or penetrant in *nr6a1a-/-* with the loss of additional *nr6a1b* alleles; this is the case for the malformations at the level of cloaca and of the pectoral fins. A dosage effect was also demonstrated previously during somitogenesis in mice (Chang et al. 2022), and is supported by the high pHaplo (0.99) and pTriplo (0.94) scores of *NR6A1*. These scores predict that the gene is likely to be sensitive to decreased DNA dosage (i.e., haploinsufficiency) and increased DNA dosage (i.e., triplosensitivity) (Collins et al. 2022). Further reports on individuals with *NR6A1* variants are warranted to elucidate genotype-phenotype correlations. Nonetheless, our results suggest that the variable expressivity of the phenotype possibly depend on NR6A1 dosage, influenced in part by the type of variant.

Amongst the clinical features, the uterine malformations were presents in three of four female individuals (MRKH syndrome in two, and hemi-uterus in one). These anomalies are suggestive of a defect in the formation or elongation process of the Müllerian ducts. Similarities in development between the oviduct in zebrafish and genital tract in mammals have been suggested. For instance, the zebrafish orthologue of *WNT4*, a well-known causal gene of MRKH syndrome, present a conserved role in gonadal ducts formation (Kossack et al. 2019; Kanamori et al. 2023). Although no obvious anomalies were observed in the gonadal duct of the 3-month-old *nr6a1a^-/-^* female zebrafish, we cannot rule out reduced penetrance of the phenotype due to the low number of surviving single *nr6a1a* homozygous. Moreover, loss of both orthologues may be required for the phenotype, that cannot be ascertained given lethality of double homozygous embryos before the onset of gonadal duct formation. However, the cloacal anomalies and defects of the terminal part of the pronephros and hindgut suggest that elongation of the oviduct might be affected in the absence of Nr6a1 protein.

A decrease in the number of ribs and ribs bearing vertebrae was observed in our zebrafish mutants, reminiscent of the defects observed in conditional knockout mice (Chang et al. 2022). Additionally, we observed a similar phenotype in one human subject, with fewer thoracic vertebrae and ribs. While 3-6% of the population may have 11 pair of ribs and vertebrae instead of 12 (Hershkovitz 2008; Yan et al. 2018; Gonzales-Portillo et al. 2021), the presence of 10 ribs is uncommon in humans. Interestingly, this reduction in the number of thoracic vertebrae and ribs might be a phenotypic clue to suspect the presence of *NR6A1* variants in humans with renal agenesis and uterine malformation.

Besides regulation of the number and segmentation of thoracolumbar vertebrae, our experiments in zebrafish supports that NR6A1 is also essential for the formation of other organs in the region of the trunk, with additional roles in kidney development and proper fusion of the pronephros and hindgut to the cloaca. The anterior shift observed in *hoxa9-13* expression may link our observations in zebrafish to the malformations observed in humans. During the formation of thoraco-lumbar somites, *Hoxa10* expression in presomitic mesoderm determines the transition between ribs-bearing and lumbar vertebrae in mice (Carapuço et al. 2005). A more rostral *HOXA10* expression, may result in a decreased number of ribs and ribs-bearing vertebrae. Moreover, precocious expression of *Hox13* paralogous in mice was shown to abruptly terminate patterning and elongation resulting in posterior truncation, and cloacal anomalies (anal atresia, anorectal agenesis or abnormal communication/septation between the bladder and the hindgut) (Young et al. 2009; van de Ven et al. 2011) and their ectopic expression causes kidney agenesis (Kmita et al. 2000) and posterior homeotic transformation of the female genital tract (Zhao and Potter 2001). Significantly, our experiments show ectopic expression of *hoxb13a* in the posterior mesoderm and endoderm in zebrafish mutants, and this may affect elongation processes in the urogenital region. Finally, in addition to posterior Hox genes, NR6A1 represses directly or indirectly other signaling molecules or transcription factors (Ibarra-Soria et al. 2023), which may also impact the development of the kidneys and uterus. Further investigation will be needed to identify the downstream targets of NR6A1 that explain the pronephros patterning defect.

In conclusion, we report *NR6A1* as a new candidate gene for renal agenesis, uterine malformation and vertebral anomalies in human. Identification of additional individuals will be necessary to confirm causality and delineate the spectrum and penetrance of anomalies associated with *NR6A1* heterozygous variants. Interestingly, the observation of cloacal malformation and pectoral fin anomalies in our zebrafish model, in addition to renal and vertebral malformations, makes *NR6A1* a compelling candidate gene for VACTERL association, in which anorectal malformation and radial ray defects are typical features. In individuals with MRKH syndrome who fulfill the criteria for an additional diagnosis of VACTERL, anorectal and renal malformations were always present, and vertebral anomalies reported in the majority of cases, highlighting the possibility of shared genetic factors and developmental pathways to explain the co-occurrence of these malformations (Bjørsum-Meyer et al. 2016). Compared to mice, zebrafish null mutants for *nr6a1a/b* exhibit later lethality in their development, allowing the investigation of late organogenesis processes. Furthermore, the survival of single *nr6a1a* homozygous mutants until the adult stage enables the observation of milder or late-onset phenotypes. In contrast, mouse models (likely through conditional knockout) will be valuable for studying the role of NR6A1 in ureteric bud branching and metanephros morphogenesis, as well as Müllerian ducts formation and patterning. Therefore, both models will complement each other in delineating further the functions of *NR6A1* in development.

## Data Availability

All data produced in the present study are available upon reasonable request to the authors

## DECLARATIONS

### Author Contribution

AJ,LF,HP,VB,BP contributed to the study conception and design. AJ,CS,AL,CP,DG,KM and VB performed clinical evaluation and clinical data collection. CS and AJ performed NGS data analyses. LF,MD,RV,HP,BP performed the in vivo and in vitro functional analyses. AJ,LF and BP wrote the first draft of the manuscript. All authors read and reviewed the manuscript.

### Ethics approval

This study was performed in line with the principles of the Declaration of Helsinki. Approval was granted by the Ethics Committee of University of British Columbia-BC Children’s and Women’s Hospital Research Ethics Board (#H09-01228); Ethical Committee of the Faculty of Medicine of the University of Liege (#2016/133) and Children’s Mercy IRB (#00000175)). All studies performed in Zebrafish were approved by the ethical committee of Liège university (protocol 2404).

### Consent to participate

Informed consent was obtained from all individual participants included in the study.

## ACKNOWLEDGEMENTS

We would like to thank the families for their participation to this study. We thank the GIGA Genomics Platform for technical assistance with NGS data generation, as well as the CARPOR Platform of the University of Liège for performing the Micro CT scans.

## SUPPLEMENTAL INFORMATION

**Supplementary Fig. 1.**
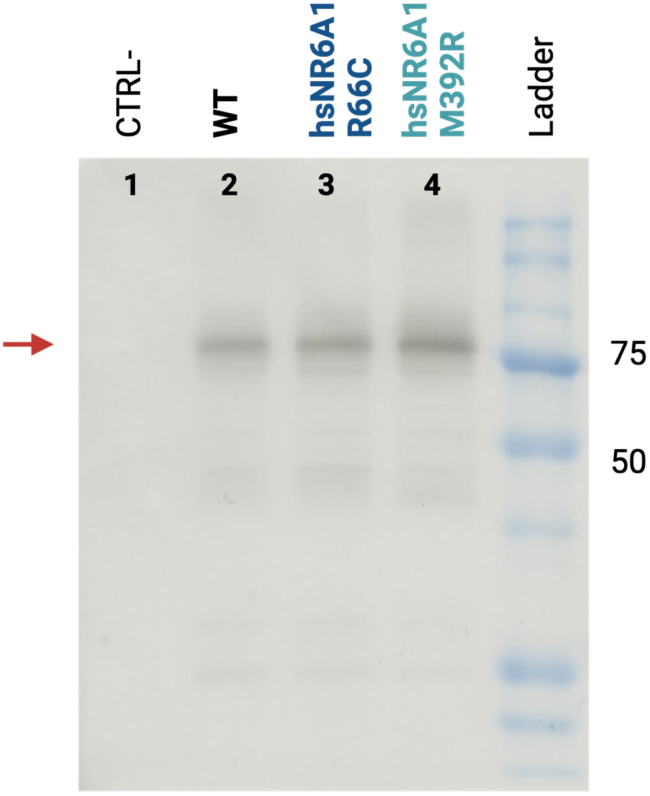
Western blot after *in vitro* translation of wildtype and mutants NR6A1. Western blot after in vitro translation showing similar concentration for the wildtype and mutants hsNR6A1. Ladder: Precision Plus Protein™ All Blue Prestained Protein Standards #1610373 Created with BioRender.com

**Supplementary Fig. 2.**
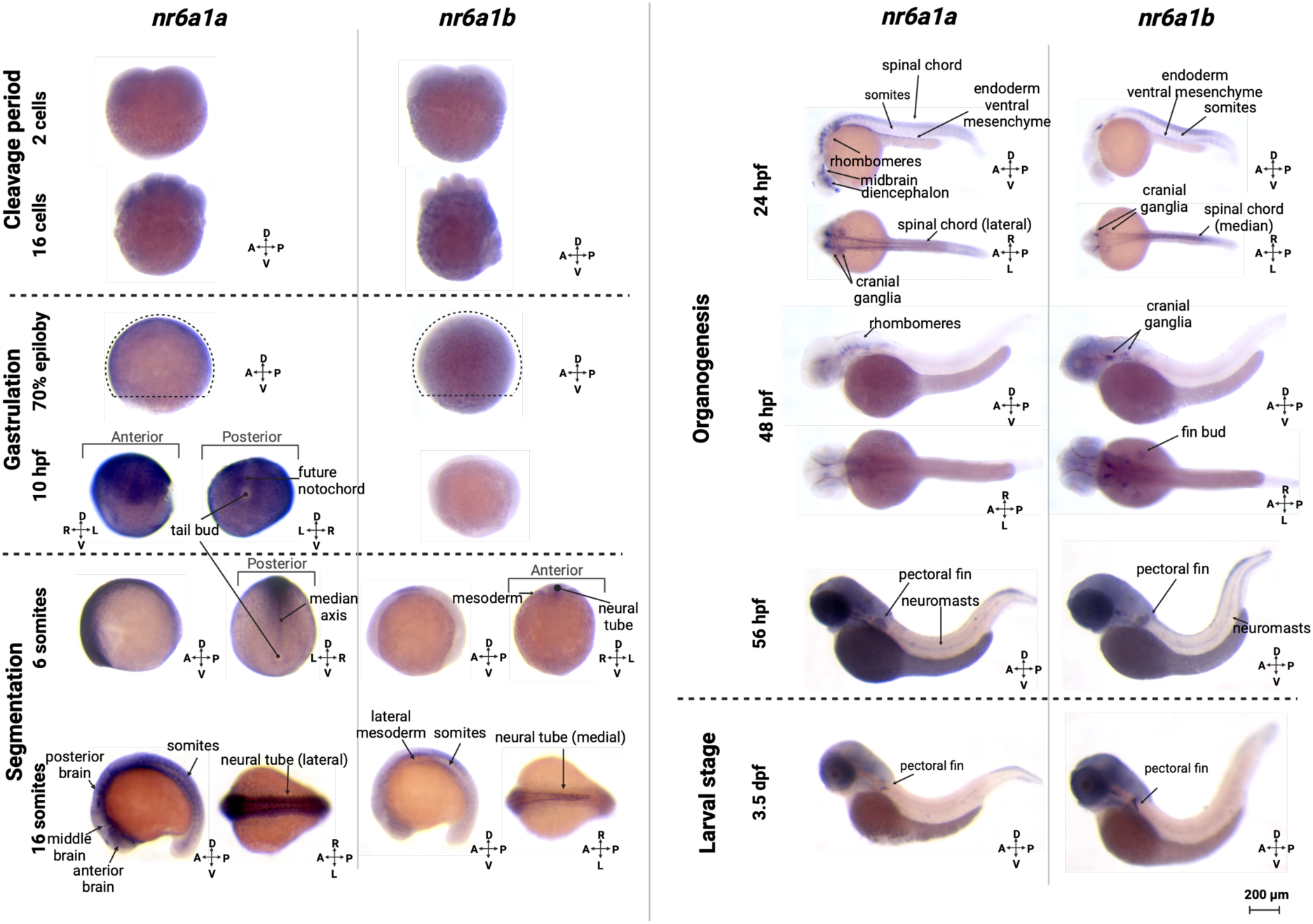
Expression of nr6a1a and nr6a1b during embryonic development. In situ hybridization at different stage of development for *nr6a1a* and *nr6a1b* on wildtype embryos. <u>Cleavage period:</u> between 0 and 2 hpf, *nr6a1a* shows a mild ubiquitous expression, this early expression (before the maternal zygotic transition) supporting maternal expression of the gene in the oocytes. In contrast, *nr6a1b* is not detected at this stage. Pictures at the 2 cells (0.75 hpf) and 16 cells (1.5hpf) stage. <u>Gastrula period:</u> At 70% epiloby (7hpf), *nr6a1a* is highly expressed ubiquitously in the gastrula while *nr6a1b* is not expressed. At 10hpf, an anteroposterior gradient is observed for *nr6a1a* with weaker signal in the posterior bud and the future notochord. The higher signal in the dorsal region is due to the higher cell mass in that region. <u>Segmentation period:</u> At the 6 somites stage (12hpf), *nr6a1a* is expressed following an anteroposterior gradient, with a stronger signal in the anterior region and absence of expression in the tailbud. At this stage, *nr6a1b* starts to be expressed in the neural tube, paraxial and lateral mesoderm. At the 16 somites stage, *nr6a1a* is still highly expressed with strong signals in the ventral region of the anterior and midbrain, as well as in the lateral part of the neural tubes and somites. *Nr6a1a* expression is lower in the posterior part of the embryo (tailbud). *Nr6a1b* is expressed in the region of the trunk in the neural tube, the somites and the lateral mesoderm. <u>Organogenesis:</u> At 24hpf, *nr6a1a* expression is restricted to the ventral part of the diencephale and midbrain, the lateral part of the rhombomeres, the spinal cord, the cranial ganglia and in the endoderm/ventral mesenchyme. A mild expression is observed in the somites. *Nr6a1b* is expressed in the cranial ganglia and the spinal cord (median region). A mild expression is detected in the endoderm/ventral mesenchyme and in the somites. At 48hpf, the expression is limited to the rhombomeres for *nr6a1a* and the cranial ganglia and fin buds for *nr6a1b*. At 56hpf and 3.5 dpf <u>(larval stage)</u>, the expression is barely detected and limited to the pectoral fin, the neuromasts and a structure (undetermined) posterior to the eyes for both genes. A=anterior; P=posterior; D=dorsal; V=ventral; R=right; L=left Created with BioRender.com

**Supplementary Fig. 3.**
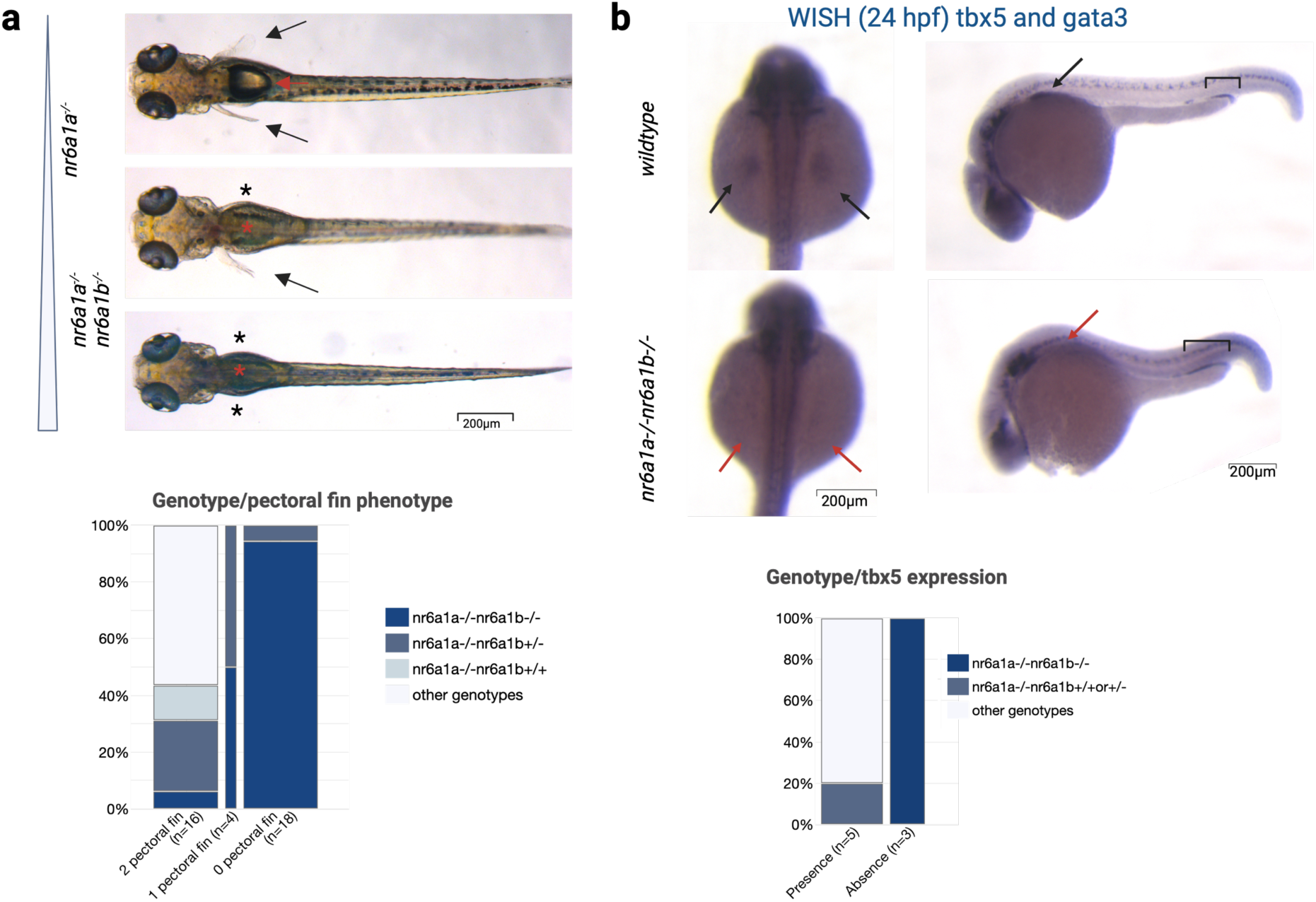
Aplasia/hypoplasia of the pectoral fins in *nr6a1a^-/-^* and *nr6a1a^-/-^ nr6a1b ^-/-^* mutants. a. Dorsal view of 5dpf larvae form *nr6a1a^+/-^nr6a1b^+/-^*incross. The pectoral fins are pointed by the black arrow (* when absent). Genotyping of 38 larvae showing that the majority of larvae without pectoral fins are double homozygous. 17/20 genotyped double homozygous larvae have no pectoral fin showing a high penetrance of the phenotype when both orthologues are knocked out. Absence of the swim bladder in affected larvae (red *) compared to the normal phenotype (red arrow). b. Whole-mount in situ hybridization for *tbx5* and *gata3* in 24hpf embryo, showing absence of *tbx5* staining and extended *gata3* staining in 18/95 embryo from *nr6a1a^+/-^ nr6a1b^+/-^* incross. Genotyping was performed for 8 embryos.

**Supplementary Fig. 4.**
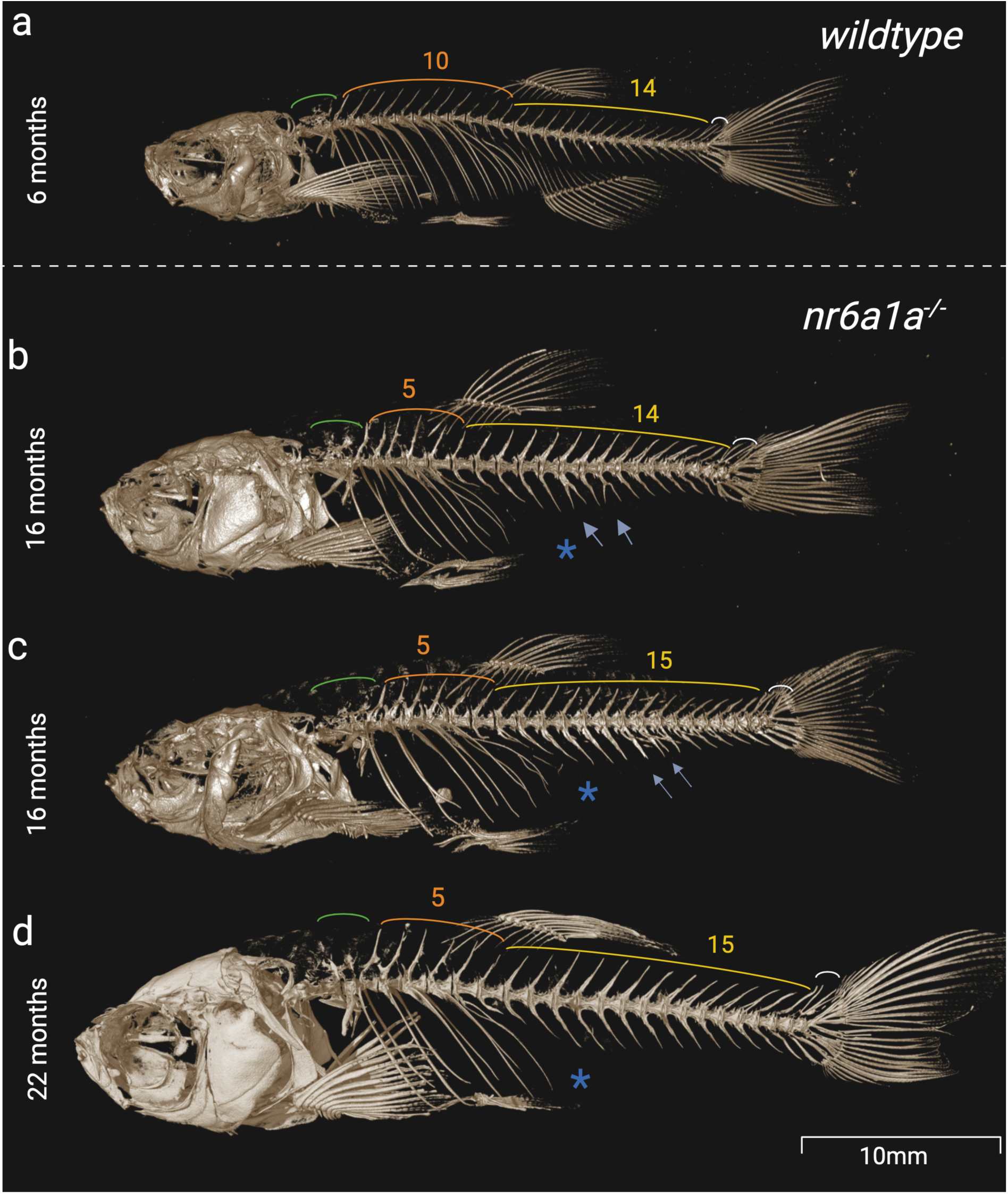
X-ray microtomography of adult wildtype and mutants. The number of rib-bearing precaudal vertebrae (orange) is decreased in the mutants (#5 instead of 10 in wildtypes). The rib-bearing precaudal vertebrae are recongizable by the presence of ribs, and the absence of hemal arches and spines. The number of caudal vertebrae (yellow, excluding the caudal fin vertebrae) varies in the mutants between 14 and 15 (number known to range from 14 to 16 in zebrafish with an average of 15 vertebrae (Bird and Mabee 2003) (Bird 2001). These vertebrae are recongnizable by the presence of hemal arches and spines. Malformed hemal arches that are unfused at the midline are observed in two mutants (b and c: blue arrow). The caudal fin vertebrae (white), that support the caudal fin rays, are normally composed of three vertebrae. The antepenultimate and penultimate vertebrae are not distinguishable in two mutants (b et d). Radials of the dorsal fin (unpaired median fin) begins between the 3rd and 5th neural spines of rib-bearing vertebrae in mutants and between the 9th and 10th neural spines of rib-bearing vertebrae in wildtype.The anal fin (beginning at the 1st caudal vertebrae in wildtype) is absent in mutants (*).The weberian vertebrae (vertebrae 1 to 4) are highlighted in green and seems normal although not investigated in details. Created with BioRender.com

**Supplementary Fig. 5.**
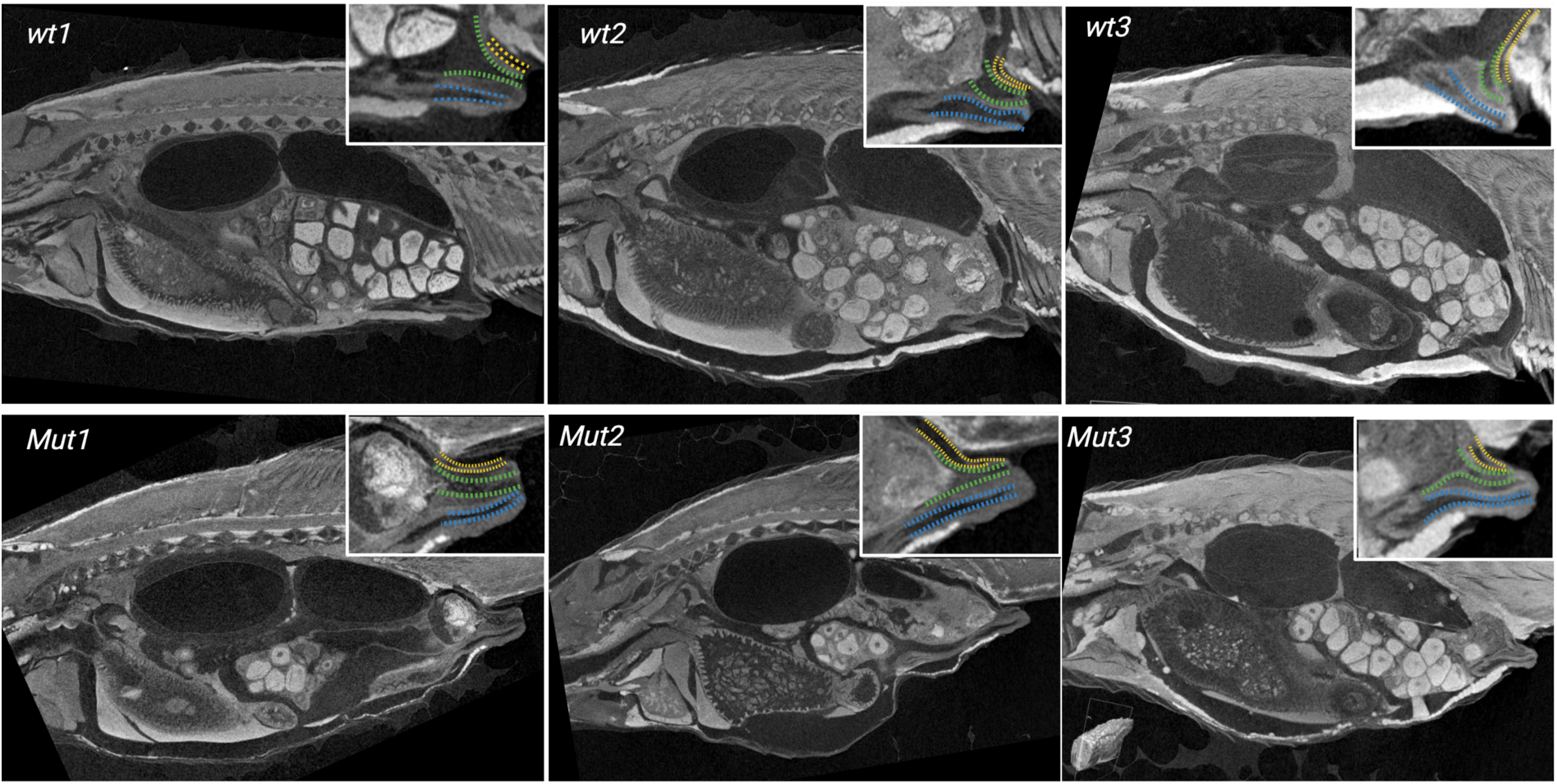
Oviduct in *nr6a1a-/-* mutants. Micro-Ct scan after Lugol staining in 3 month-old wildtype (n=3) and *nr6a1a^-/-^* mutants (n=3).In mutants, the oviduct seems formed. The duct seems obstructed by degenerating oocytes.Created with BioRender.com

**Supplementary Fig. 6.**
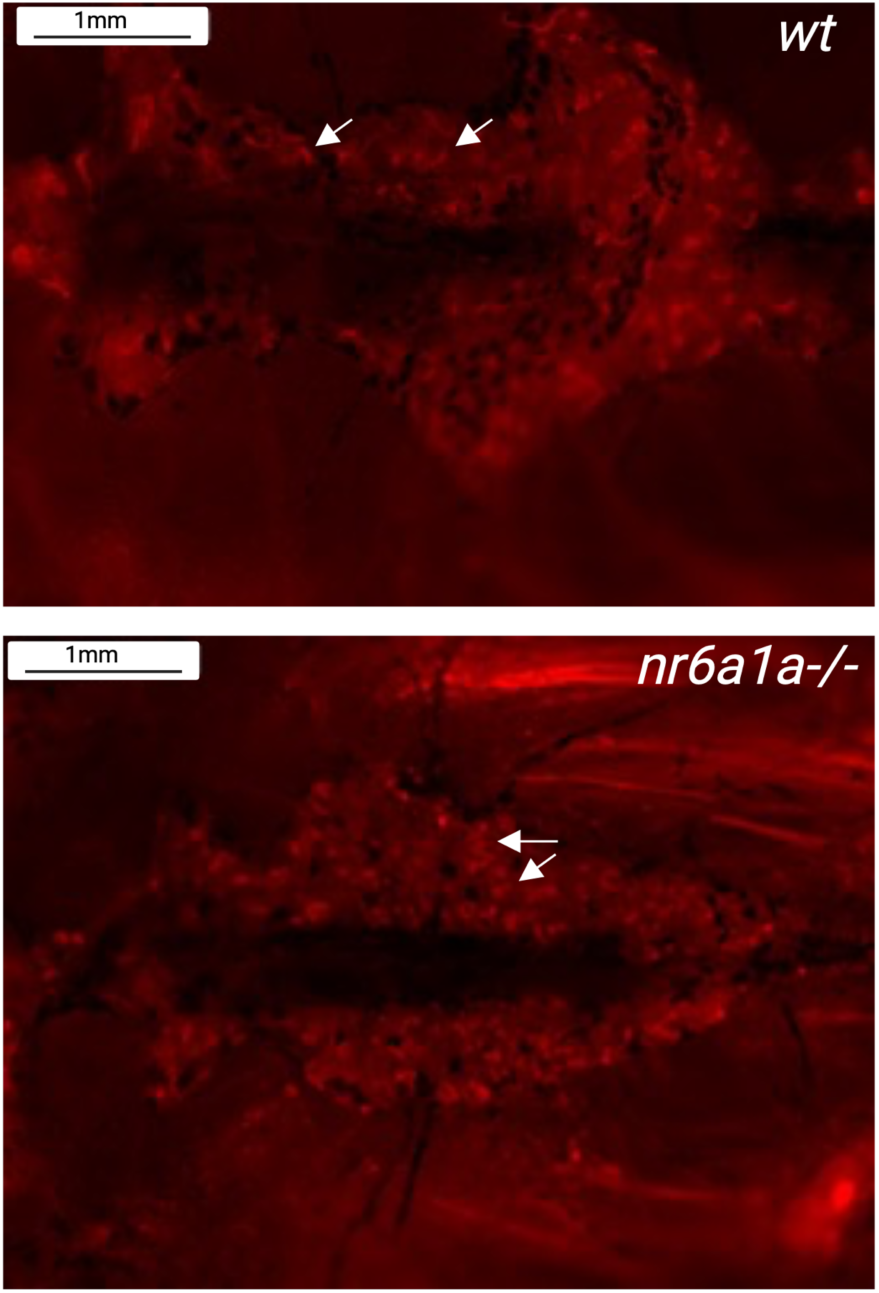
Uptake of Rhodamine-Dextran seems normal in *nr6a1a^-/-^* mutants. Rhodamine-dextran compounds seem normally incorporated by the epithelial cells in the proximal circumvulated tubule (red fluorescence, black arrow). Created with BioRender.com

**Supplementary Fig. 7.**
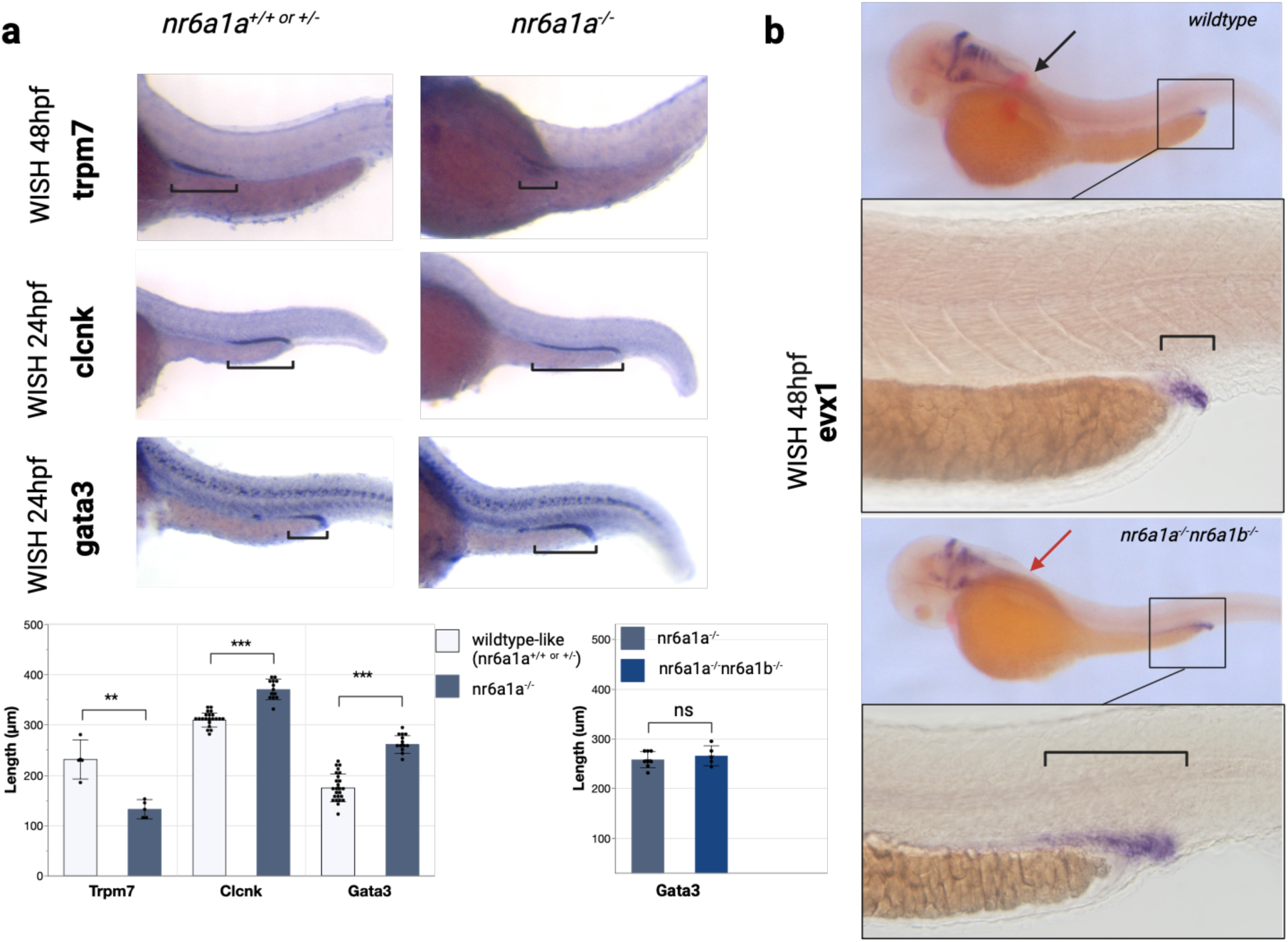
Pronephros expression. a. Expression of segment markers was performed on embryos from *nr6a1a^+/-^* incross (*clcnk*) or *nr6a1a^+/-^ nr6a1b^+/-^* incross (*trpm7, gata3*) at 24hpf (*gata3, clcnk*), 48hpf (*trpm7*). Measurement (in μm) were statistically different between wildtypes and *nr6a1a^-/-^* for *trpm7* (n=8), *clnck* (n=33) and *gata3* (n=38). No statistically significant difference was observed for *gata3* between *nr6a1a^-/-^* (n=8) and double mutants (n=5). b. Expression of *evx1* (coloration black NBT-BCIP) and *tbx5* (coloration fast-red) was performed on 89 embryos from *nr6a1a^+/-^nr6a1b^+/-^* incross at 48 hpf. 8/89 embryos show no *tbx5* staining (previously shown to be associated with the mutant phenotype) and marked anterior extension of the *evx1* staining. Lateral view of the embryos. Brackets indicate the expression domain in each embryo. Created with BioRender.com

**Supplementary Fig. 8.**
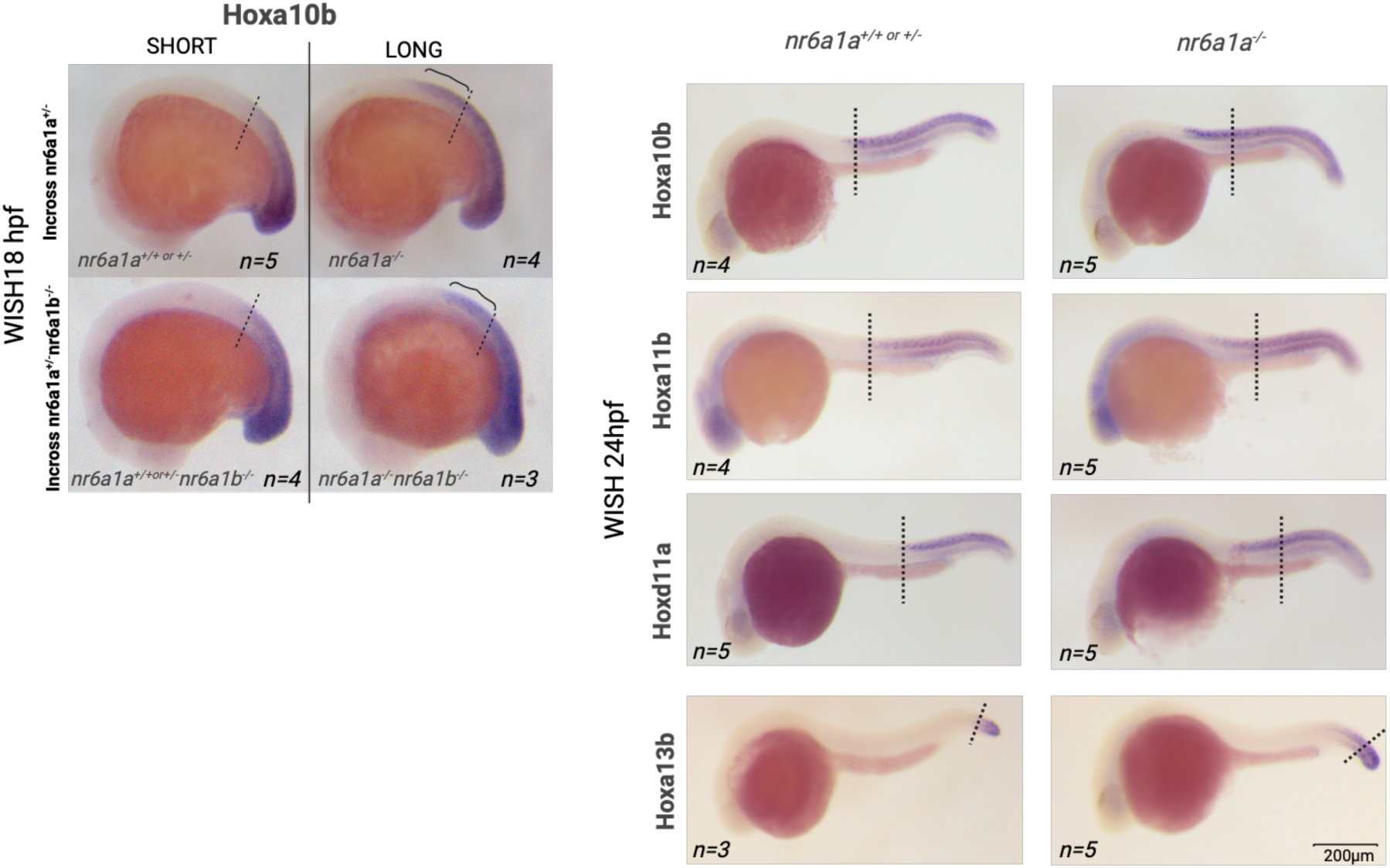
Abnormal patterning of posterior Hox in *nr6a1a^-/-^* and *nr6a1a ^-/-^ nr6a1b ^-/-^* mutants. a. Comparison of whole-mount is situ hybridization with *hoxa10b* at 18hpf in embryos from incross between *nr6a1a^+/-^nr6a1b^+/+^*and embryo from an incross between *nr6a1a^+/-^nr6a1^-/-^*. No significant difference is observed between *nr6a1b^-/-^* and wildtypes (SHORT phenotype). Extended anterior staining (LONG phenotype) in simple *nr6a1a^-/-^* and double *nr6a1a^-/-^nr6a1b^-/-^*mutants. b. Whole-mount is situ hybridization with *hoxa10b, hoxa11b, hoxd11a, hoxa13b* at 24hpf in embryos from an incross between *nr6a1a^+/-^nr6a1b^-/-^* showing an extended anterior staining in *nr6a1a^-/-^* mutants. Created with BioRender.com

